# Racialized experience, biomarkers of lead exposure, and later-life cognition: a mediation analysis

**DOI:** 10.1101/2023.04.22.23288920

**Authors:** Tara E. Jenson, Kelly M. Bakulski, Linda Wesp, Keith Dookeran, Ira Driscoll, Amy E. Kalkbrenner

## Abstract

We evaluated the role of the neurotoxicant lead (Pb) in mediating racial disparities in later-life cognition in 1,085 non-Hispanic Black and 2,839 non-Hispanic white participants in NHANES (1999-2002, 2011-2014) 60+ years of age. We operationalized Black race as a marker for the experience of racialization and exposure to systemic racism. We estimated patella bone Pb via predictive models using blood Pb and demographics. Concurrent cognition (processing speed, sustained attention, working memory) was measured by the Digit Symbol Substitution Test (DSST) and a global measure combining four cognitive tests. To obtain the portion mediated, we used regression coefficients (race on Pb * Pb on cognitive score)/(race on cognitive score), adjusting for age, NHANES cycle, and sample weights. Other confounder adjustment (education, poverty income ratio, smoking) was limited to the mediator-outcome (i.e., Pb-cognition) pathway because these factors do not lie upstream of race and so cannot confound associations with race. Pb was estimated to mediate 0.6% of the association between race and global cognition, and 4% of the DSST. Our results suggest that later-life cognitive health disparities may be impacted by avoidable lead exposure driven by environmental injustice, noting that a large proportion of the pathway of systemic racism harming cognition remains.

## Introduction

Some age-related decline in later-life cognitive function is expected in humans (1,2). However, more precipitous, abnormal decline in cognitive function can occur due to disease conditions like dementia or exposure to neurotoxicants (1,2). Later-life cognitive impairment is an important marker of progression toward Alzheimer’s Disease (AD) and other dementias (3).

Disparities in later-life cognitive function and higher dementia burden among Black versus White Americans are well-documented and persist even after accounting for other risk factors for AD that include early life family circumstances, educational attainment, occupation, and income level (4,5,6,7). The reasons for this disparity are likely multifactorial and related to the embodiment of entrenched and systemic racialized experiences that Black communities face in the United States (6,7,8,9,10).

Higher levels of lead (Pb) exposure in Black communities as a factor for racial disparities in later-life cognitive function has yet to be investigated (11). Pb is a bioaccumulative neurotoxicant with adverse effects on cognitive development and function from childhood through mid-life even at low levels of exposure (12,13,14,15). The etiologic window encompassing initiation of physiological processes and accumulation of damage leading to abnormal cognitive decline is thought to span years to decades (2,12). Epidemiological evidence consistently and robustly links environmental Pb to accelerated cognitive aging, poorer later-life cognitive function, AD diagnosis and AD-related mortality (12,16,17,18,19). Due to historic and continuing trends of systemic environmental injustice and segregation in the U.S. driving poverty and deteriorating housing and water infrastructure, racial disparities of higher yet avoidable Pb exposure have been tracked since the 1970s and persist currently (11). Yet no study to date has explored levels of Pb exposure and later-life cognitive outcomes among Black populations.

We conducted a mediation analysis to evaluate the proportion of later-life cognitive deficit among people who identify as Black, whereby Black race is considered a marker of racialized experience and exposure to systemic racism (10), that may be due to higher exposure to environmental Pb. To assess the proportion of the disparity mediated by environmental Pb exposure independently of other influences, and to minimize bias due to overadjustment for variables we argue cannot be confounders of race (20), we performed a mediation analysis where only the regression pathway between Pb and cognitive performance was fully adjusted for socioeconomic and behavior confounders. Identifying modifiable pathways related to prevention or reduction of adverse health outcomes is a key goal of public health research (21). Understanding the extent to which avoidable Pb exposure may explain later-life cognitive health disparities is an important step towards achieving this goal.

## Methods

### Study population

The sample was comprised of 3,924 individuals participating in the United States National Health and Nutrition and Examination Survey (NHANES; cycles 1999-2002 (N=2,224) and 2011-2014 (N=1,700) (Web Figure 1)). NHANES is an ongoing, annual, cross-sectional study that uses a stratified multi-stage probability sampling design to capture a representative sample of the US population while over-sampling for adults 60+ years old and African Americans (22). We included participants 1) aged 60 years or older (an eligibility criteria for the cognitive assessments), 2) randomly subsampled to provide blood specimen collection for the metals assay, 3) identified as non-Hispanic Black or non-Hispanic White, and 4) who underwent cognitive assessment.

**Figure 1.**
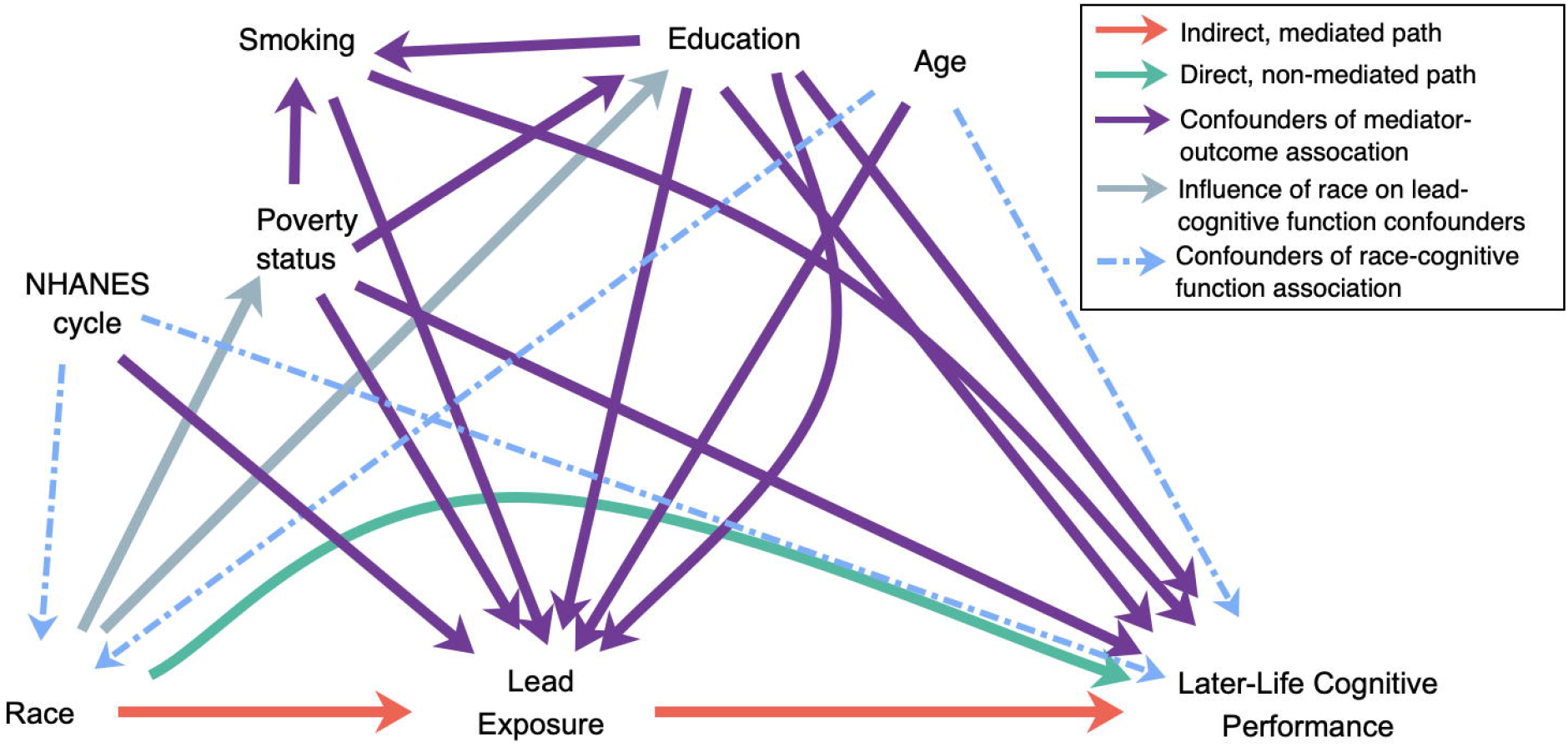
Directed acyclic graph (DAG) causal diagram representing hypothesized causal relationships among mediators and confounders of the association between marker of racism/racialized experience (Black vs White), lead exposure, and later-life cognitive function.

Participants provided written, informed consent at the time of data collection (22), and the current analyses were conducted under the oversight of the Institutional Review Board (IRB) of the University of Wisconsin – Milwaukee IRB #22.298-UWM.

### Cognitive assessment

During the 2011-2014 NHANES cycles, four different cognitive tests were administered by trained NHANES interviewers during an in-person visit in the Mobile Examination Center (MEC) (23): i) Consortium to Establish a Registry for Alzheimer’s Disease (CERAD) immediate word learning (score 0-10) ii) CERAD delayed recall word learning (score 0-10), iii) Animal Fluency test (score 0-40), and iv) the Digit Symbol Substitution test (DSST) (score 0-130), a component of the Wechsler Adult Intelligence Scale (23). The DSST measures processing speed, sustained attention, and working memory (executive function). Collectively, the four tests assess multiple domains of cognitive function including memory, executive function, sustained attention, and working memory (23). For this analysis, all four scores for each participant were standardized into z-scores (mean 0, standard deviation 1), averaged, then re-standardized into a final z-score. The tests were conducted in written or spoken English, Spanish, Korean, Vietnamese, traditional or simplified Mandarin, or Cantonese (23). Scores were missing for about 2% due to not consenting to recorded session, unspecified communication problem, physical limitation, refusal, or equipment failure (23).

For the 1999-2002 cycles, only the DSST was administered during the household interview portion of the survey in spoken and written English (24). Scores were missing for about 8% due to: inability to read or write, home environment preventing completion of assessment, physical limitations, cognitive limitations, or assertion of not enough time to complete assessment (24).

### Race

Our main independent variable of interest was self-reported race (non-Hispanic Black or non-Hispanic White) (25). We considered race to be a socially constructed marker encompassing the joint effects of a range of characteristics and racialized experiences that may include but are not limited to physical phenotype, parental physical phenotype, cultural context, childhood and neighborhood socioeconomic status, and experience of racism and discrimination (10,20,26). We hypothesized that in the context of our analysis and causal interpretation, effects of race and cumulative lifetime Pb exposure causally/temporally occur upstream from our outcome of later-life (age 60 and older) cognitive function.

### Lead measures

The mediator of interest in our analyses was patella bone lead (Pb) as bone serves as a repository of cumulative Pb exposure across the lifespan (12,27). To estimate patella Pb, we used NHANES blood Pb measures. A random half of participants aged 12 and older were eligible for collection of whole blood specimens for the 2013-2014 cycles, while all participants aged one year and older were eligible for whole blood sampling for the 1999-2002 and 2011-2012 cycles (28,29,30,31). Whole blood samples were collected by venipuncture (32), frozen (-20°C for 1999-2002 cycles, -30°C for the 2011-2012 cycles, and -70°C for 2013-2014 cycles) and shipped at cold temperatures (2-8°C) to the Division of Environmental Health Laboratory Sciences, National Center for Environmental Health, Centers for Disease Control and Prevention for analysis (28,29,30,31). For 1999-2002 cycles, whole blood samples were diluted and blood Pb concentrations were quantified from liquid samples using a PerkinElmer Model SIMAA 6000 simultaneous multi-element atomic absorption spectrometer with Zeeman background correction by procedures described elsewhere (28,29). For 2011-2014 cycles, whole blood samples were diluted and treated with anticoagulants, then assayed for Pb content by inductively coupled plasma mass spectrometer by procedures described elsewhere (30,31). The lower limit of detection (LLOD) for 1999-2002 samples was determined by NHANES laboratory procedures as 0.05 µg/dL (28,29). The LLODs for 2011-2012 and 2013-2014 samples determined by NHANES laboratory procedures were 0.25 µg/dL and 0.10 ug/dL, respectively (30,31). For all cycles, Pb assay results determined to be below the LLOD were assigned as per NHANES laboratory procedures values of the respective cycle LLOD divided by √2.

We used measures of blood Pb, demographic and health measures to estimate patella bone Pb levels using prediction models developed and validated by Park et al. (27). We selected patella bone models over tibia due to higher validation performance, and we combined the 14-predictor and 6-predictor models given the variable measures available across NHANES cycles (27). Web Table 1 shows the model and predictor variables used to impute patella Pb concentrations and includes continuous measures of blood Pb as described above, continuous body-mass index (kg/m^2^) measured during MEC visit, and the following self-reported information collected during in-home interviews: continuous age, three-category education level (grade 11 or less, high school graduate/GED or some college, or college graduate or higher) and smoking status coded as described below (33,34,35).

**Table 1.**
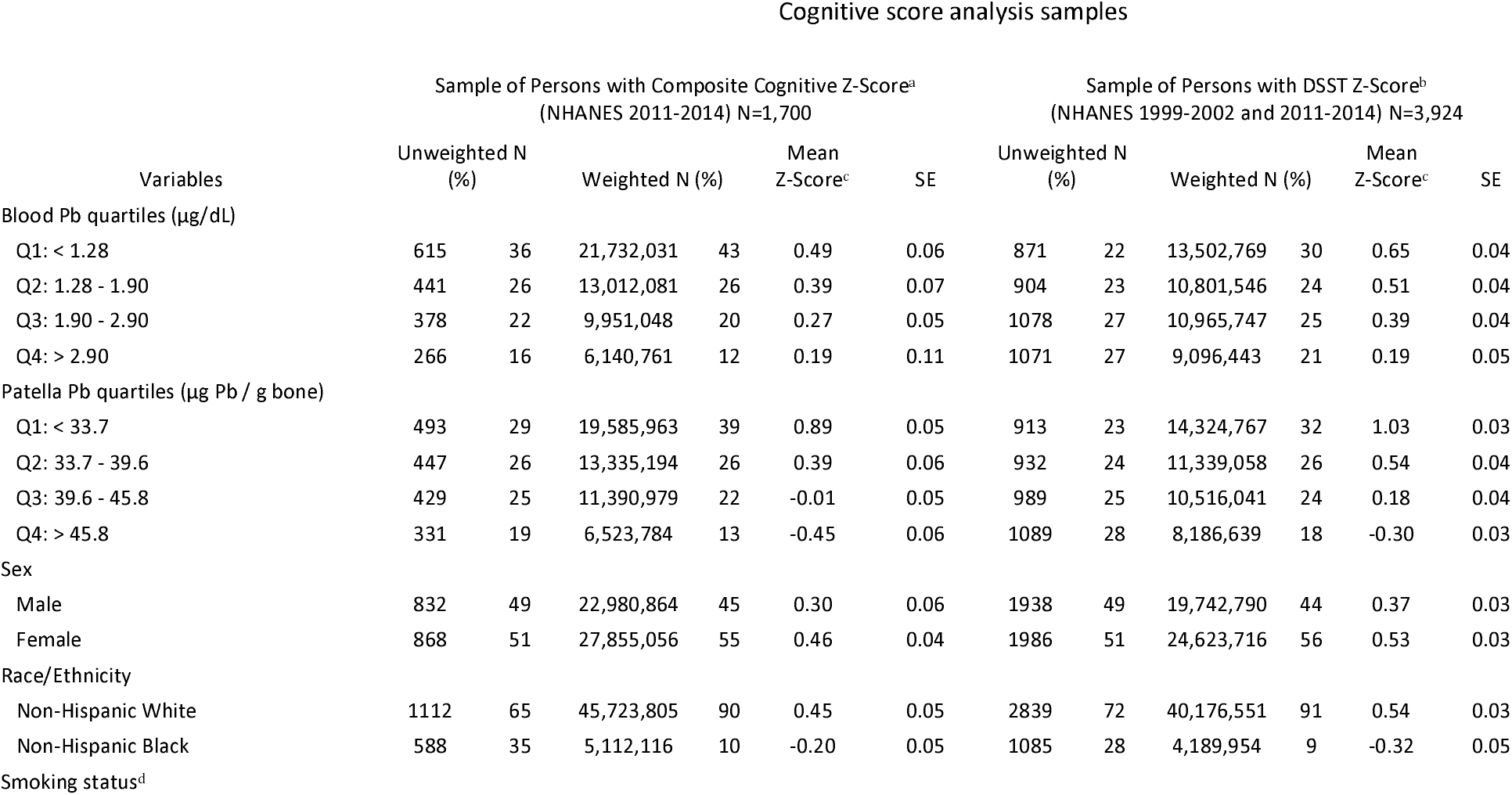
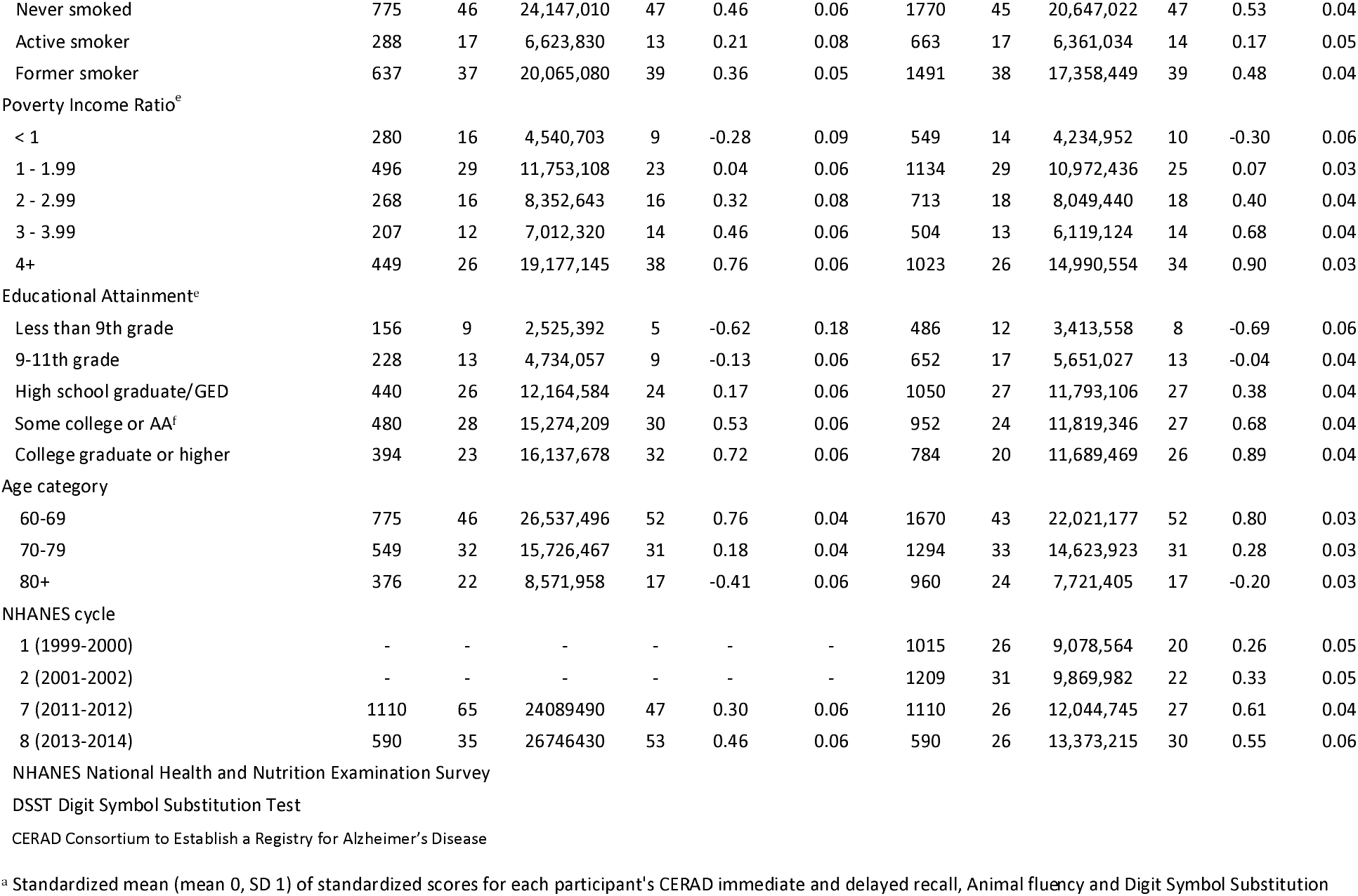
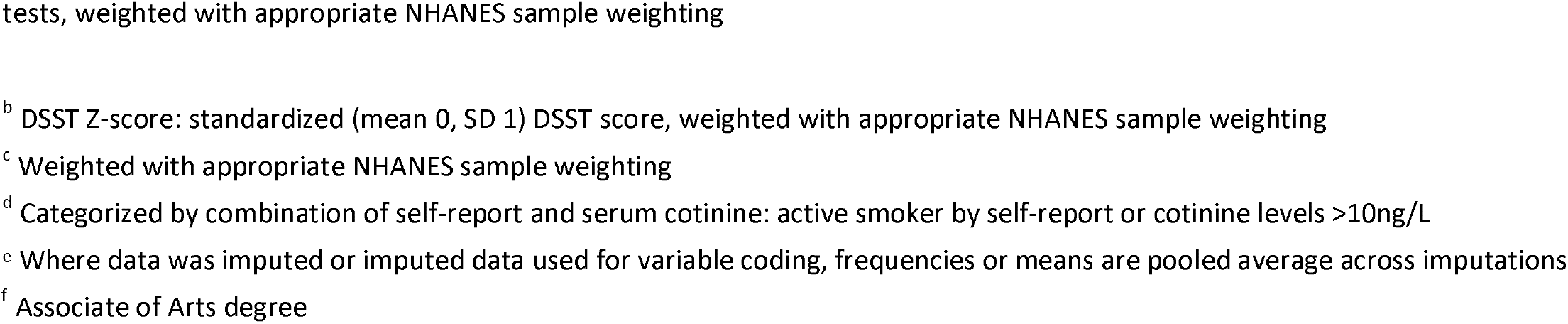
Characteristics of NHANES participants 60+ years and older sub-sampled for blood metals, for composite cognitive z-score analysis (2011-2014) and Digit Symbol Substitution z-score analysis (1999-2002 and 2011-2014).

We used continuous measures of patella Pb in our regression models based on better model fit with cognitive measures as indicated by lower Akaike Information Criteria (AIC) over other transformations and categorizations.

### Selection and measurement of other characteristics

We identified potential confounders for our analysis by a priori assumptions of association of these covariates with the direct and indirect pathways (3,4,5,6,13,14,15,16,17). We constructed a directed acyclic graph (Figure 1) describing the hypothesized causal associations to construct a “minimally sufficient” set of confounders (17). Adjusting for educational attainment and poverty status in our models as confounders allowed us to block backdoor paths to later-life cognitive function through occupation. Final set of covariates included NHANES cycle (defined as each two-year survey period) and the following variables measured by self-report during in-home interviews: continuous age (in years), educational attainment (less than 9th grade, 9-11th grade, high school graduate/GED, some college or Associate of Arts, college graduate or higher), ratio of family income to poverty guidelines (range of 0 to 5), and smoking status coded as never, former or active smoker.

We defined active smokers as those who self-reported via in-home interview as current cigarette smokers or had serum cotinine measures >10ng/mL (35,36,37,38). NHANES serum cotinine assays were conducted using isotope dilution-high performance liquid chromatography/atmospheric pressure chemical ionization tandem mass spectrometry (ID HPLC-APCI MS/MS). The LLOD for 1999-2000 samples was 0.05 ng/mL and that of 2001-2002 and 2011-2014 samples was 0.015 ng/mL, with those below the LLOD assigned as per NHANES laboratory procedures values of the respective cycle LLOD divided by √2 (28,29,37,38).

### Statistical approach

We used SAS 9.4 via SAS OnDemand for Academics (SAS Institute, Cary, NC), with SURVEYREG packages used to appropriately account for the complex, multistage, probability sampling design used for collection of NHANES data (39). We incorporated MEC survey weights applicable for participants sampled for blood specimen collection constructed for combined NHANES cycles as directed (40).

We evaluated whether our associations differed by NHANES cycle and time. This was important because of study design differences and temporal trends in Pb exposures (41) and social perceptions of race. In all cases cross-product terms between NHANES cycle with Pb and with race yielded p values > 0.05, supporting combining NHANES cycles.

### Approach for missing data

To avoid potential bias due to dropping about 21% of people missing one or more variables, we considered multiple imputation. We determined covariate and cognitive score outcomes were missing completely at random or missing at random, using “RBTest” in the “RBTest” (42) R package (R version 4.2.1) (43). These patterns are appropriate for remediation with multiple imputation to fill in missing information and include these persons in the models. We conducted multiple imputation using SAS PROC MI with fully conditional specification to create five imputed datasets from the larger superset of all individuals 60 years and older in NHANES cycles 1999-2002 and 2011-2014, using a broad set of predictors: cognitive score (outcome), race (independent variable), blood Pb levels (mediator of interest), confounders identified above, sex, marital status, body mass index, and systolic and diastolic blood pressure. Regression analyses were conducted using SAS PROC MIANALYZE on imputed data to pool association and variance estimates. (See Web Tables 2a, 2b, 3a, and 3b for description of missingness).

**Table 2.** Characteristics of NHANES participants 60+ years and older sub-sampled for blood metals, by continuous variable measures, NHANES 1999-2002 and 2011-2014.

|  | Composite Cognitive Z-Score<br>Analysis Sample<br>(NHANES 2011-2014)<sup>a</sup> |  | DSST Z-Score Analysis Sample<br>(NHANES 1999-2002 and 2011-2014)<sup>b</sup> |  |
| --- | --- | --- | --- | --- |
|  | Mean<sup>c</sup> | SE<sup>c</sup> | Mean<sup>c</sup> | SE<sup>c</sup> |
| Age | 69.7 | 0.30 | 70.4 | 0.21 |
| Blood lead (µg/dL) | 1.83 | 0.08 | 2.19 | 0.06 |
| Patella lead (µg Pb / g bone) | 36.6 | 0.41 | 38.2 | 0.29 |
| CERAD immediate recall |  |  |  |  |
| Raw score | 7.7 | 0.09 | - | - |
| Z-score<sup>d</sup> | 0.21 | 0.05 | - | - |
| CERAD delayed recall |  |  |  |  |
| Raw score | 6.1 | 0.09 | - | - |
| Z-score<sup>d</sup> | 0.18 | 0.04 | - | - |
| Animal Fluency |  |  |  |  |
| Raw score | 18.1 | 0.22 | - | - |
| Z-score<sup>d</sup> | 0.36 | 0.04 | - | - |
| DSST |  |  |  |  |
| Raw score | 52.1 | 0.68 | 49.8 | 0.53 |
| Z-score<sup>d</sup> | 0.47 | 0.04 | 0.46 | 0.03 |
| Composite z-score<sup>d</sup> | 0.39 | 0.04 | - | - |

NHANES National Health and Nutrition Examination Survey
DSST Digit Symbol Substitution Test
<sup>a</sup> Composite Cognitive z-score sample: unweighted N=1,700; weighted N = 50,835,920
<sup>b</sup> DSST z-score sample: unweighted N=3,924; weighted N = 44,366,505
<sup>c</sup> Means and SEs weighted for NHANES survey weighting
<sup>d</sup> Standardized score, mean of 0 and standard deviation of 1

### Mediation analyses

We used decomposition mediation analysis to assess the proportion of racial disparity in later-life cognitive performance mediated by environmental Pb exposure independently of other influences. We used separate models to examine separate cognitive outcomes of both DSST score and composite cognitive score. We assert that socioeconomic factors are not acting as confounders of race, but instead are themselves affected by the multiplicity of life experience encompassed by race (gray arrows, Figure 1). Adjusting any part of the mediation pathway where race is acting as predictor for these socioeconomic factors risks over-adjustment bias (20). Thus, only the association between Pb exposure and cognitive function should be adjusted for education, poverty status, and smoking status (purple arrows, Figure 1). We have dubbed this approach of a fine-tuned, “just right” confounder adjustment for each pathway of the mediation analysis our “Goldilocks Scenario” (in reference to the protagonist of “Goldilocks and the Three Bears” (44)). All mediation pathways were adjusted for age as a confounder of race due to different proportions of participation by race over time, and NHANES cycle to adjust for potential differences in social norms and study procedures between cycles (blue dotted arrows, Figure 1).

We used the step-by-step mediation methods described by Baron and Kenny (Web Figure 2) (45,46). Other all-inclusive mediation software packages like R mediate and SAS CAUSALMED do not provide a mechanism for differential confounder adjustment on separate parts of the mediation pathway (47). In the step-by-step Baron and Kenny approach, the mediated or indirect association *a*b* can be calculated as a product of the regression estimates for a) the exposure -> mediator pathway, and b) the mediator -> outcome pathway. The portion of the total association of exposure on outcome through the mediated pathway then is the quantity *a*b*.

**Figure 2.**
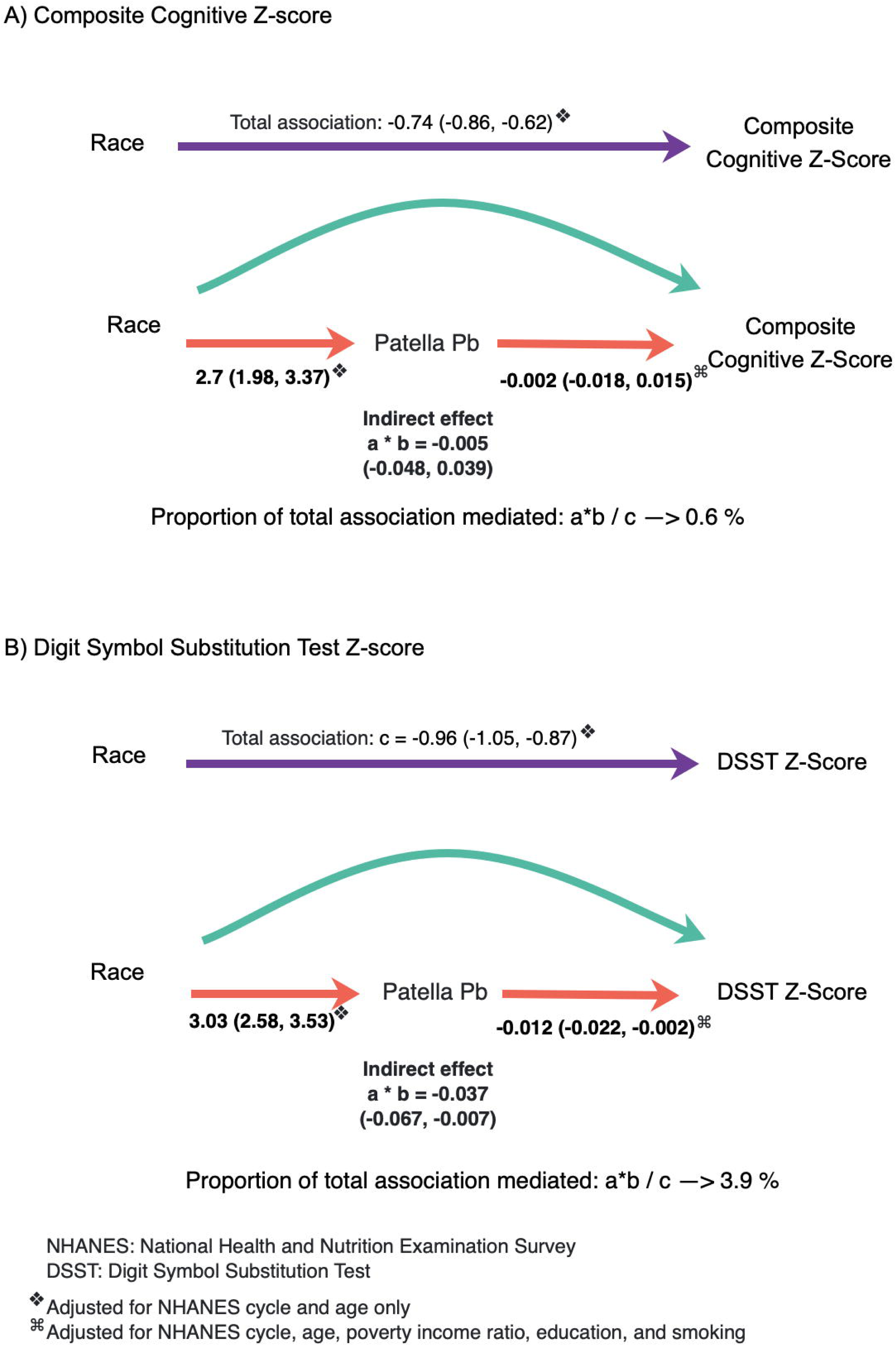
Associations between race (Black vs. White), patella lead, and cognitive score 60+ years and older participants, NHANES 1999-2002 and 2011-2014

### Precision estimates for mediation analysis

We calculated confidence intervals (CIs) around the mediated effect (indirect association) using methods described and validated by others (48). This approach entails using the estimates (b_a_ and b_b_) and standard errors (SE_a_ and SE_b_) generated via PROC SURVEYREG and PROC MIANALYZE for the a and b pathway regressions to calculate a combined mediation estimate standard error SE_ab_ as shown in equation 1.

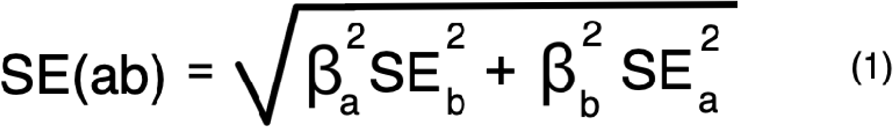

We then calculated 95% CIs as b_ab_ +/-1.96*(SE_ab_) (48,49).

### Comparison analyses

To contrast our just-right, “Goldilocks Scenario 1” of differential confounding adjustment along each of the mediation pathways with standard mediation approaches, we conducted a minimally adjusted mediation model (Scenario 2) where all regression pathways were adjusted only for age and NHANES cycle and an over-adjusted mediation model (Scenario 3) additionally adjusted for education level, poverty status, and smoking status on all pathways.

### Sensitivity analyses

We repeated the three mediation scenarios described above (“Goldilocks” just-right adjusted Scenario 1, under-adjusted Scenario 2, and over-adjusted Scenario 3) using SAS PROC CAUSALMED and GENMOD to compare our results with those arising from the mediation packages. CAUSALMED and GENMOD models did not account for survey design as they do not currently support accounting for these elements.

## Results

### Descriptive Statistics

Our combined 1999-2002 and 2011-2014 sample had a mean age of 70 years and was predominantly White (90-91%) (Table 1). The 1999-2002 sample overall had lower levels of educational attainment than the 2011-2014 subsample. Cognitive scores were lower for earlier NHANES cycles, Black persons, active smokers, older individuals, and those with lower education and income (Table 1). Mean blood and patella Pb concentrations for the combined sample were higher (2.19 μg/dL and 38.2 μg Pb/g bone, respectively) compared to the later cycles (1.83 μg/dL and 36.6 μg Pb/g bone) (Table 2). Levels of Pb in both samples were along expected demographic patterns, with higher Pb for persons of older age, lower educational attainment, lower income, and identifying as Black (Table 3).

**Table 3.** Characteristics of NHANES participants 60+ years old, mean measured blood lead and estimated patella lead, for composite cognitive z-score analysis (2011-2014) and Digit Symbol Substitution Test z-score analysis (1999-2002 and 2011-2014).

| Variables | Sample of Persons with Composite Cognitive Z-Score<sup>a</sup><br>(NHANES 2011-2014) N=1,700 |  |  |  | Sample of Persons with DSST Z-Score<sup>b</sup><br>(NHANES 1999-2002 + 2011-2014) N=3,924 |  |  |  |
| --- | --- | --- | --- | --- | --- | --- | --- | --- |
|  | Blood Pb<br>(µg/dL) |  | Patella Pb<sup>c</sup><br>(µg Pb / g bone) |  | Blood Pb<br>(µg/dL) |  | Patella Pb<sup>c</sup><br>(µg Pb / g bone) |  |
|  | Mean<sup>d</sup> | SE | Mean<sup>d</sup> | SE | Mean<sup>d</sup> | SE | Mean<sup>d</sup> | SE |
| Sex |  |  |  |  |  |  |  |  |
| Male | 2.2 | 0.1 | 36.6 | 0.6 | 2.6 | 0.1 | 38.2 | 0.4 |
| Female | 1.6 | 0.1 | 36.7 | 0.4 | 1.9 | 0.0 | 38.2 | 0.3 |
| Race/Ethnicity |  |  |  |  |  |  |  |  |
| Non-Hispanic White | 1.8 | 0.1 | 36.5 | 0.4 | 2.1 | 0.1 | 38.0 | 0.3 |
| Non-Hispanic Black | 2.2 | 0.1 | 38.0 | 0.5 | 2.6 | 0.1 | 39.7 | 0.4 |
| Smoking status<sup>e</sup> |  |  |  |  |  |  |  |  |
| Never smoked | 1.5 | 0.1 | 34.9 | 0.5 | 1.8 | 0.0 | 36.9 | 0.4 |
| Active smoker | 2.5 | 0.2 | 36.8 | 0.6 | 2.9 | 0.1 | 38.3 | 0.4 |
| Former smoker | 2.0 | 0.2 | 38.6 | 0.6 | 2.4 | 0.1 | 39.7 | 0.4 |
| Poverty Income Ratio |  |  |  |  |  |  |  |  |
| < 1 | 2.1 | 0.2 | 40.7 | 0.6 | 2.6 | 0.1 | 42.5 | 0.4 |
| 1 - 1.99 | 2.0 | 0.1 | 40.1 | 0.6 | 2.3 | 0.1 | 41.7 | 0.4 |
| 2 - 2.99 | 1.8 | 0.1 | 38.0 | 0.5 | 2.2 | 0.1 | 39.4 | 0.3 |
| 3 - 3.99 | 1.7 | 0.2 | 36.4 | 0.9 | 2.1 | 0.1 | 37.5 | 0.6 |
| 4+ | 1.7 | 0.1 | 33.0 | 0.6 | 2.0 | 0.1 | 34.1 | 0.4 |
| Educational Attainment |  |  |  |  |  |  |  |  |
| Less than 9th grade | 2.3 | 0.2 | 45.4 | 0.8 | 2.9 | 0.1 | 46.5 | 0.5 |
| 9-11th grade | 1.8 | 0.1 | 43.0 | 0.5 | 2.4 | 0.1 | 43.8 | 0.3 |
| High school graduate/GED | 1.9 | 0.2 | 38.0 | 0.5 | 2.2 | 0.1 | 38.9 | 0.3 |
| Some college or AA<sup>f</sup> | 1.8 | 0.1 | 37.2 | 0.5 | 2.0 | 0.1 | 38.0 | 0.4 |
| College graduate or higher | 1.8 | 0.1 | 31.9 | 0.6 | 2.0 | 0.1 | 32.5 | 0.4 |
| Age category |  |  |  |  |  |  |  |  |
| 60-69 | 1.8 | 0.1 | 31.4 | 0.3 | 2.1 | 0.1 | 32.5 | 0.3 |
| 70-79 | 1.8 | 0.1 | 40.6 | 0.4 | 2.2 | 0.1 | 41.7 | 0.2 |
| 80+ | 2.0 | 0.1 | 45.5 | 0.3 | 2.4 | 0.1 | 47.7 | 0.3 |
| NHANES cycle |  |  |  |  |  |  |  |  |
| 1 (1999-2000) | - | - | - | - | 2.8 | 0.1 | 40.4 | 0.4 |
| 2 (2001-2002) | - | - | - | - | 2.5 | 0.1 | 40.2 | 0.4 |
| 7 (2011-2012) | 1.9 | 0.1 | 37.0 | 0.5 | 1.9 | 0.1 | 37.0 | 0.5 |
| 8 (2013-2014) | 1.7 | 0.1 | 36.3 | 0.6 | 1.7 | 0.1 | 36.3 | 0.6 |

NHANES: National Health and Nutrition Examination Survey
DSST: Digit Symbol Substitution Test
<sup>a</sup> Adults 60+ in 2011-2014 cycles identifying as non-Hispanic Black or non-Hispanic White, sampled for blood metals assay
<sup>b</sup> Adults 60+ 1999-2002 and in 2011-2014 cycles identifying as non-Hispanic Black or non-Hispanic White, sampled for blood metals assay
<sup>c</sup> Calculated using prediction models developed by Park et al. 2009 incorporating blood Pb measure, age, education level, smoking status, and body mass index
<sup>d</sup> Weighted with appropriate NHANES sample weighting
<sup>e</sup> Categorized by combination of self-report and serum cotinine: active smoker by self-report or cotinine levels >10ng/L
<sup>f</sup> Associate of Arts degree

### Pathway Results

The overall association of race and cognitive scores showed 74% (95% CI -0.86, -0.62) to 96% (95% CI -1.05, -0.87) standard deviation lower scores for composite and DSST cognitive scores, respectively among Black compared to White participants (Table 4).

**Table 4.** Associations between race (Black vs. White) and composite cognitive z-score (NHANES 2011-2014) and Digit Symbol Substitution z-score (60 years and older participants, NHANES 1999-2002 and 2011-2014).

| Model | Composite cognitive Z-score<sup>a</sup><br>(NHANES 2011-2014) |  |  | DSST Z-Score<sup>b</sup><br>(NHANES 1999-2002 + 2011-2014) |  |  |
| --- | --- | --- | --- | --- | --- | --- |
| | $\beta$ | 95%<br>Lower CI | 95%<br>Upper CI | $\beta$ | 95%<br>Lower CI | 95%<br>Upper CI |
| Model 1<sup>c</sup> | -0.74 | -0.86 | -0.62 | -0.96 | -1.05 | -0.87 |

NHANES: National Health and Nutrition Examination Survey
DSST: Digit Symbol Substitution Test
CERAD: Consortium to Establish a Registry for Alzheimer's Disease
<sup>a</sup> Standardized mean (mean 0, SD 1) of standardized scores for each participant's CERAD, immediate and delayed recall, Animal fluency and DSST, weighted with appropriate NHANES sample weighting
<sup>b</sup> DSST Z-score: standardized (mean 0, SD 1) DSST score, weighted with appropriate NHANES sample weighting
<sup>c</sup> Model 1: adjusted for NHANES cycle and age only

On separate parts of the mediation pathways, Black participants (vs. Whites) had higher Pb levels: 2.7 μg Pb per g bone (95% CI 2.0, 3.4) in the later cycles and 3.0 μg Pb per g bone (95% CI 2.5, 3.5) in the combined sample (path a, Model 1 estimates, Table 5). Composite cognitive z-score was 0.002 lower for each unit increase in Pb (95% CI ⍰0.018, 0.015). DSST z-score decreased by 0.012 (95% CI ⍰0.022, -0.002) for each unit increase in Pb (path b, Model 1 estimates, Table 5).

**Table 5.** Individual pathway associations for step-by-step mediation analysis of race (Black vs. White) and cognitive scores, mediated by patella lead measure (60+ years and older participants, NHANES 1999-2002 and 2011-2014)

| Path | Composite cognitive Z-score<sup>a</sup><br>(NHANES 2011-2014) |  |  |  |  |  | Digit Symbol Substitution Test Z-Score<sup>b</sup><br>(NHANES 1999-2002 + 2011-2014) |  |  |  |  |  |
| --- | --- | --- | --- | --- | --- | --- | --- | --- | --- | --- | --- | --- |
|  | Model 1<sup>c</sup> |  |  | Model 2<sup>d</sup> |  |  | Model 1<sup>c</sup> |  |  | Model 2<sup>d</sup> |  |  |
| | $\beta$ | 95%<br>Lower CI | 95%<br>Upper CI | $\beta$ | 95%<br>Lower CI | 95%<br>Upper CI | $\beta$ | 95%<br>Lower CI | 95%<br>Upper CI | $\beta$ | 95%<br>Lower CI | 95%<br>Upper CI |
| a<sup>e</sup> | 2.68 | 1.98 | 3.37 | 0.96 | 0.58 | 1.35 | 3.03 | 2.54 | 3.53 | 1.22 | 0.93 | 1.51 |
| b<sup>f</sup> | -0.04 | -0.05 | -0.02 | 0.00 | -0.02 | 0.01 | -0.05 | -0.06 | -0.04 | -0.01 | -0.02 | 0.00 |
| c<sup>g</sup> | -0.74 | -0.86 | -0.62 | -0.52 | -0.62 | -0.42 | -0.96 | -1.05 | -0.87 | -0.68 | -0.74 | -0.61 |

NHANES: National Health and Nutrition Examination Survey
DSST: Digit Symbol Substitution Test
CERAD: Consortium to Establish a Registry for Alzheimer's Disease
<sup>a</sup> Standardized mean (mean 0, SD 1) of standardized scores for each participant's CERAD, immediate and delayed recall, Animal fluency and DSST, weighted with appropriate NHANES sample weighting
<sup>b</sup> Digit Symbol Substitution Test (DSST) Z-score: standardized (mean 0, SD 1) DSST score, weighted with appropriate NHANES sample weighting
<sup>c</sup> Model 1: adjusted for NHANES cycle and age only
<sup>d</sup> Model 2: adjusted for NHANES cycle, age, poverty income ratio, education level, and smoking status
<sup>e</sup> Path a: Association between race (Black vs white) and patella lead ( $\mu\text{g Pb / g bone}$ )
<sup>f</sup> Path b: Association between patella lead ( $\mu\text{g Pb / g bone}$ ) and cognitive score, adjusted for race
<sup>g</sup> Path c: Total association between race (Black vs white) and cognitive z-score

### Mediation Results

Under our recommended “Goldilocks” Scenario adjusted “just-right” for confounders (rather than over or under-adjusted), out of a total association of 74% lower composite cognitive score for Black participants (vs. Whites), we estimated Pb to mediate 0.6% (Table 6; Figure 2A). Out of a total association of 96% lower DSST score for Black participants (vs. Whites), we estimated Pb to mediate 4% (Table 6; Figure 2B).

**Table 6.**
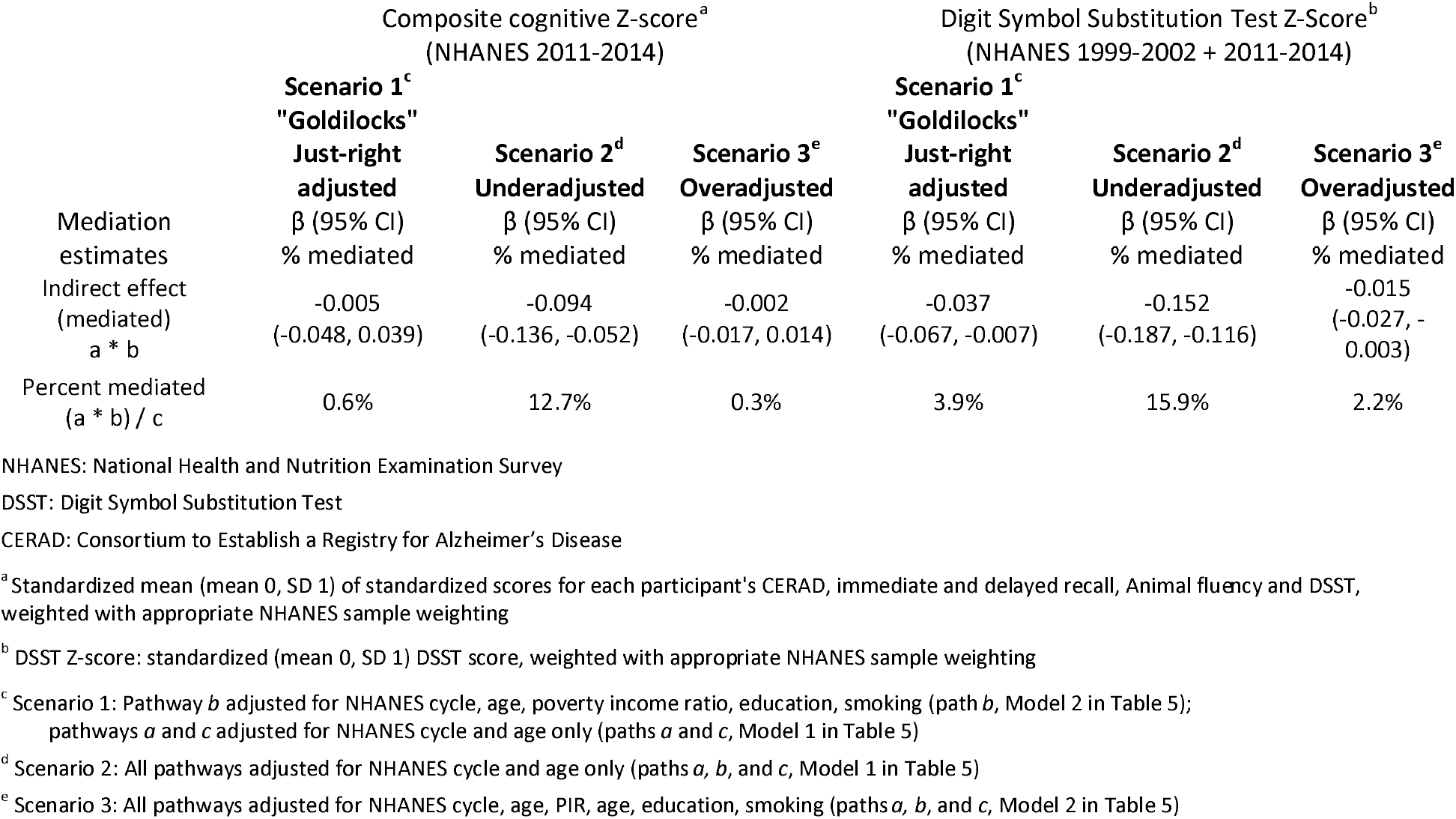
Proportion of racial disparity in composite cognitive z-score and Digit Symbol Substitution Test z-score mediated by patella lead (60+ years and older participants, NHANES 1999-2002 and 2011-2014)

### Sensitivity Analyses

In sensitivity analyses, the over-adjusted Scenario 3 attenuated the indirect association through patella Pb to 0.3% (beta -0.002; 95% CI -0.017, 0.014) of the total association of race with composite cognitive z-score. The under-adjusted Scenario 2 increased this estimate to 13% mediated through Pb (beta -0.094; 95% CI -0.136, -0.052) (Table 6). For DSST z-scores, the over-adjusted Scenario 3 attenuated the indirect association of patella Pb to 2% (beta -0.015; 95% CI ⍰0.027, ⍰0.003), while the under-adjusted Scenario 2 increased this to 16% (beta ⍰0.152; 95% CI ⍰0.187, -0.116) (Table 6).

Sensitivity analyses conducted using SAS PROC CAUSALMED and GENMOD (without survey weighting) produced similar mediation estimates for each of the adjustment scenarios as our primary analyses (Web Tables 4 and 5).

## Discussion

Racial disparities in health in the US are systemic and on-going, with mechanisms only partly understood. Whether higher exposures to environmental neurotoxicants account for some portion of these disparities has been largely unexplored. We found no examples of studies assessing mediation of racial disparities in later-life cognition by either environmental exposures broadly or Pb exposure specifically. Our unique contribution is in demonstrating that in a large United States representative sample, 4% of the racial disparity between older Black and White adults in cognition – specifically processing speed, sustained attention, and working memory, was mediated through cumulative lead exposure.

Others have explored mediation of racial disparities in later-life cognition by various socioeconomic, health, and psychosocial factors including educational attainment, early life family circumstances, income, wealth, occupation, sibling loss, and stressors across the life course. Individually these factors are estimated to account for less than 20% of racial disparities in later-life cognitive function (7,50,51,52), excepting educational attainment which may account for up to 35% of these disparities (6).

Our mediation question was built upon strong evidence of association for the individual pathways between race, lead exposure and later-life cognitive function, also demonstrated in our analyses. We report cognitive deficits for Black participants compared to their White counterparts, consistent with the literature (6,7,8,50,51,52). Our results confirm racial environmental injustice evidenced by higher Pb exposure among Black older adults also demonstrated by others (53,54). Our results also support the link of Pb with cognitive domains of processing speed, sustained attention, and working memory (DSST) and to a lesser degree for composite cognitive score. We note that published evidence suggests strong links between Pb and various measures of later-life cognitive performance, such as with cognitive decline (16,18,19,55), eye-hand coordination (56), executive functioning, and verbal memory and learning (56,57). In bringing these individual pathways together in mediation analysis, we demonstrated that 4% of racial disparities in DSST score was mediated through higher patella Pb among Black participants vs. White participants.

Given our findings of mediation of racial differences through Pb exposure for DSST score but not composite z-score, perhaps brain regions subserving specific domains of cognition measured by the DSST (processing speed, sustained attention, and working memory) are more sensitive to Pb exposure compared to composite z-score as a global measure of cognition. An alternate explanation is that neurological impacts of Pb are more observable for the higher ranges of Pb exposure present for the study population in the DSST analyses, which included earlier NHANES cycles with higher historical Pb levels.

A strength of our study was separately investigating DSST, measured across four cycles of NHANES vs. the other composite z-score components conducted across only two cycles. This increased sample size and reflected earlier time periods with higher Pb exposures. Furthermore, the DSST, with a possible but rarely achieved score of 130, is less subject to ceiling effects than the composite z-score CERAD recall components that range from 0-10. Ceiling effects are a type of measurement error where the upper score limit is artificial, not allowing observation of a hypothetical true possible score beyond the upper limit (58). Not accounting for cognitive testing ceiling effects is likely to attenuate an observed association. (59,60).

A key innovation of our approach is in handling a special case of mediation where socioeconomic factors are adjusted for only on the mediator-outcome pathway (our “Goldilocks” just-right approach). This allowed a more fine-tuned estimate of the mediated path that minimized over-adjustment for socioeconomic factors we hypothesized are themselves influenced by race as a marker of racism and thus cannot be confounders of race. While this approach requires a step-by-step analysis and does not lend itself to all-inclusive mediation statistics packages, our sensitivity analyses using PROC GENMOD and CAUSALMED demonstrated that our estimates and confidence intervals were validly calculated. Our approach also allowed us to use PROC SURVEYREG to correctly incorporate NHANES complex survey design features.

Our nuanced “Goldilocks” approach generated a slightly different mediated proportion (4%) compared with alternate standard approaches explored in our sensitivity analyses that we argued are less valid. Not adjusting for confounding influences of socioeconomics on Pb exposure yielded a proportion mediated of around 16%, while over-adjusting race for socioeconomics yielded a proportion mediated around 2%. It is possible other mediation questions could be better illuminated by carefully considering adjustment at each leg of the pathways, as we have done.

A further strength of our study is deployment of patella Pb as a measure of long-term Pb exposure, allowing us to estimate 10-15 years of cumulative Pb exposure, longer than the 30-day half-life of blood Pb measures alone (12,27). A limitation of this measure is that it is unlikely to reflect childhood or early adult lead exposure. Underestimation of our proportion mediated (4%) may also be caused by measurement error in Pb. Lead exposure is neurotoxic across the lifespan, and Pb exposure during fetal development, childhood, and beyond, likely influences later-life cognition to some degree. Furthermore, the originators of the patella Pb prediction models note that their prediction equations (developed using samples from older males) may underestimate Pb levels in older females due to differences in bone remodeling rates (27).

Limitations of our study include having cognitive measures from only a single time point for each participant, differential item functioning in cognitive measures used, survival bias in our sampling, and residual confounding. Repeat cognitive measures would better characterize a trajectory of change in cognitive function with aging. “Differential item functioning” (DIF) in cognitive measures used may have introduced measurement error into our data due to racial disparities in cognitive performance based on differences in sociocultural norms and experiences associated with test performance (58,61,62,63); our study methods did not include DIF detection procedures for assessing bias in cognitive measures by racial grouping. Survival bias is a concern when studying racial disparities in later-life health outcomes given Black populations in the US have overall shorter life expectancies than Whites, which may have led to underestimating the role of Pb in mediating association of race and later-life cognition in our analyses (58,64). Residual confounding may be present in our analyses due to available socioeconomic measures limited to years of education and family income. Neither of these measures reflect education quality or accessibility, wealth attainment, or inherited wealth, all of which are important in considering the historic and continuing effects of systemic, structural racism on education and wealth in the US (62,65).

## Conclusion

Our findings suggest a relatively small proportion of racial disparities in later-life cognition may be mediated through cumulative Pb exposure. However, even in providing a small contribution to cognitive disparities, Pb as a modifiable risk factor may have a significant impact on the overall prevention and costs associated with later-life cognitive deficits and dementia. Our “Goldilocks” approach to mediation analysis could have great public health utility in further understanding underlying pathways of racial health inequalities. In addition, many of the limitations here may have led to an underestimation of the racial disparity itself or the proportion mediated, so that the true impacts may be larger. A preponderance of research demonstrates that there are no safe levels of Pb exposure nor any stages of life during which adverse effects of Pb are negligible; thus, we should require no further evidence to enact policies and interventions that minimize environmental Pb exposure for all populations and groups, especially marginalized groups at higher risk of these exposures due to the legacy of structural racism.

## Supporting information

Web Material

## Data Availability

All data produced in the present study are available upon reasonable request to the authors

## Abbreviations

AD: Alzheimer’s Disease
Cd: Cadmium
CERAD: Consortium to Establish a Registry for Alzheimer’s Disease
DIF: Differential Item Functioning
DSST: Digit Symbol Substitution Test
LLOD: lower limit of detection
MEC: Mobile Examination Center
NHANES: National Health and Nutrition Examination Survey
Pb: Lead

## Acknowledgements

This work was supported by the National Center for Advancing Translational Sciences, National Institutes of Health (award TL1TR001437); and the National Institutes of Health (grant R01 AG070897). The content is solely the responsibility of the authors and does not necessarily represent the official views of the NIH.

