## Supplementary material for "Racialized experience, biomarkers of lead exposure, and later-life cognition: a mediation analysis": Web Material

Co-authors

Tara E. Jenson, Kelly M. Bakulski, Linda Wesp, Keith Dookeran, Ira Driscoll, Amy E. Kalkbrenner

Contents

Web Figure 1. NHANES sample selection for individuals 60 years and older with blood lead measures eligible for cognitive assessment testing (page 2)

Web Table 1. Linear prediction model for imputing patella lead based on blood lead levels and key demographic and health measures (page 3)

Web Table 2a. Observation dropout due to combined cognitive score and covariate missingness for NHANES participants 60+ years and older sub-sampled for blood metals, for composite cognitive z-score analysis (2011-2014, N = 1700) (page 4)

Web Table 2b. Covariate missingness for NHANES participants 60+ years and older sub-sampled for blood metals, for composite cognitive z-score analysis (2011-2014, N = 1700) (page 5)

Web Table 3a. Observation dropout due to combined cognitive score and covariate missingness for NHANES participants 60+ years and older sub-sampled for blood metals, for DSST z-score analysis (NHANES 1999-2002 and 2011-2014, N=3,924) (page 6)

Web Table 3b. Covariate missingness for NHANES participants 60+ years and older sub-sampled for blood metals, for Digit Symbol Substitution Test z-score analysis (1999-2002 and 2011-2014, N = 3924) (page 7)

Web Figure 2. Baron and Kenny approach for step-by-step estimation of mediated effects

Directed acyclic graph (page 8)

Web Table 4. Sensitivity Analyses – mediation results using PROC GENMOD and CAUSALMED for composite cognitive score analyses (NHANES 2011-2014) (page 9)

Web Table 5. Sensitivity Analyses – mediation results using PROC GENMOD and CAUSALMED for DSST z-score analyses (NHANES 1999-2002 + 2011-2014) (page 10)

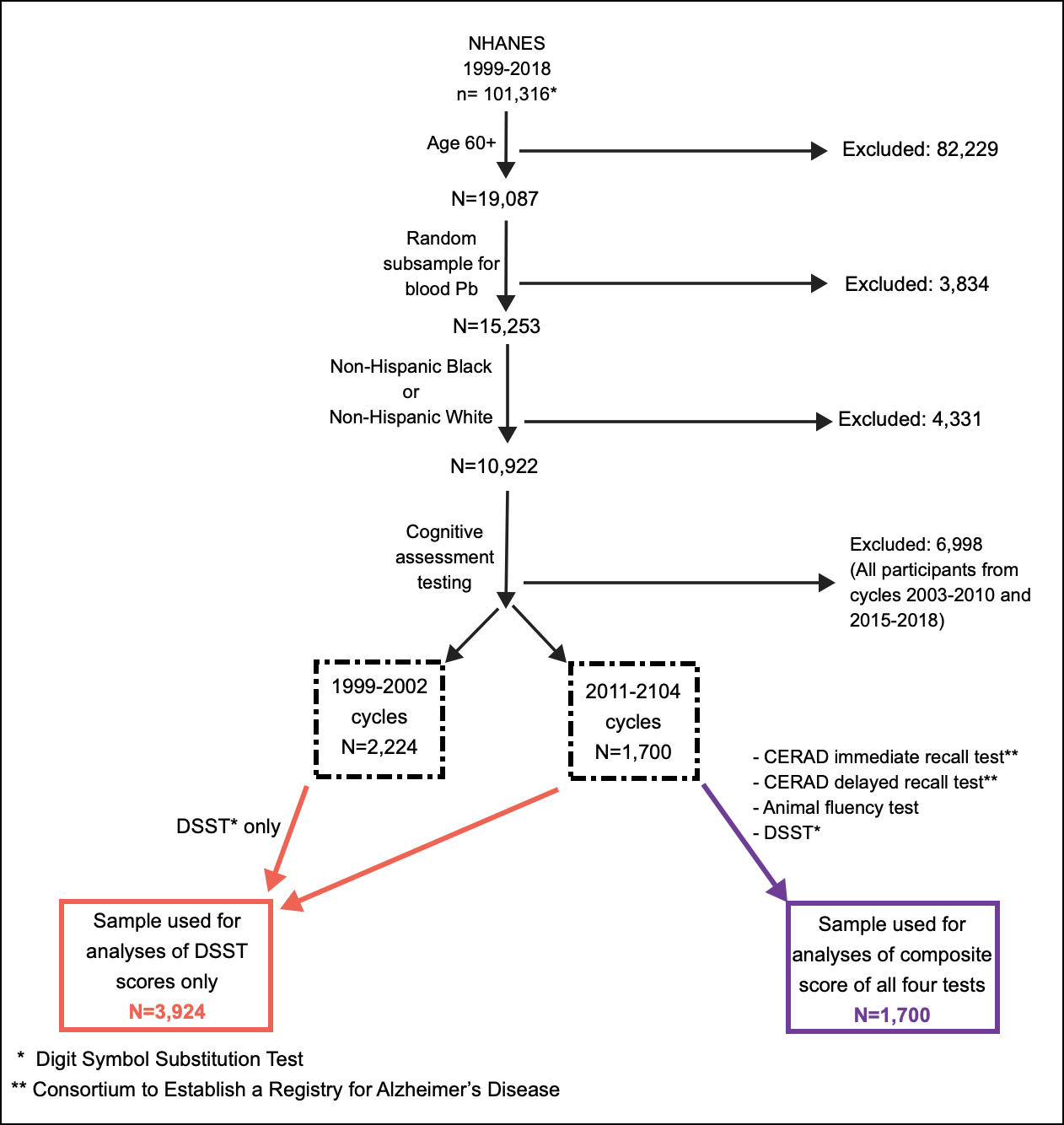

*Web Figure 1.* NHANES 1999-2018: Sample selection for individuals 60 years and older with blood lead measures who underwent cognitive assessment testing

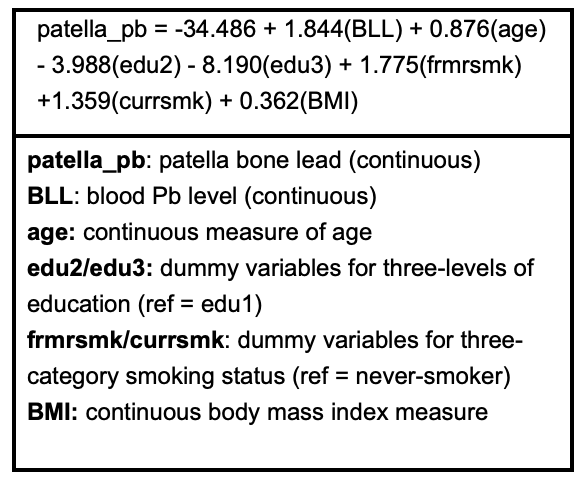

*Web Table 1.* Linear prediction model for imputing patella lead based on blood lead levels and key demographic and health measures

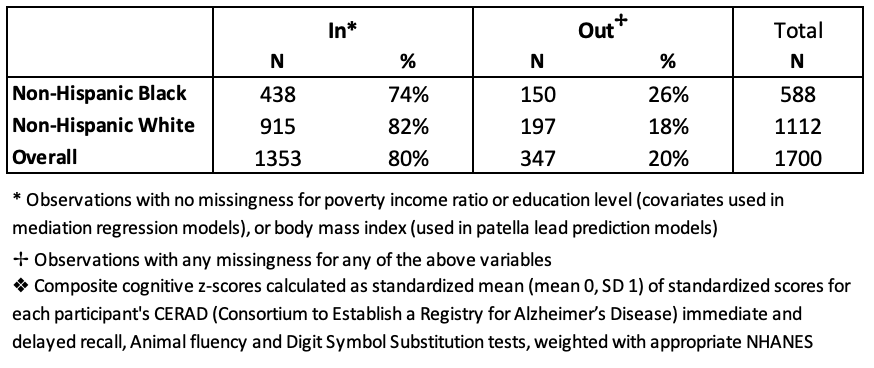

*Web Table 2a*. Observation dropout due to combined cognitive score and covariate missingness for NHANES participants 60+ years and older sub-sampled for blood metals, for composite cognitive z-score analysis (2011-2014, N = 1700)

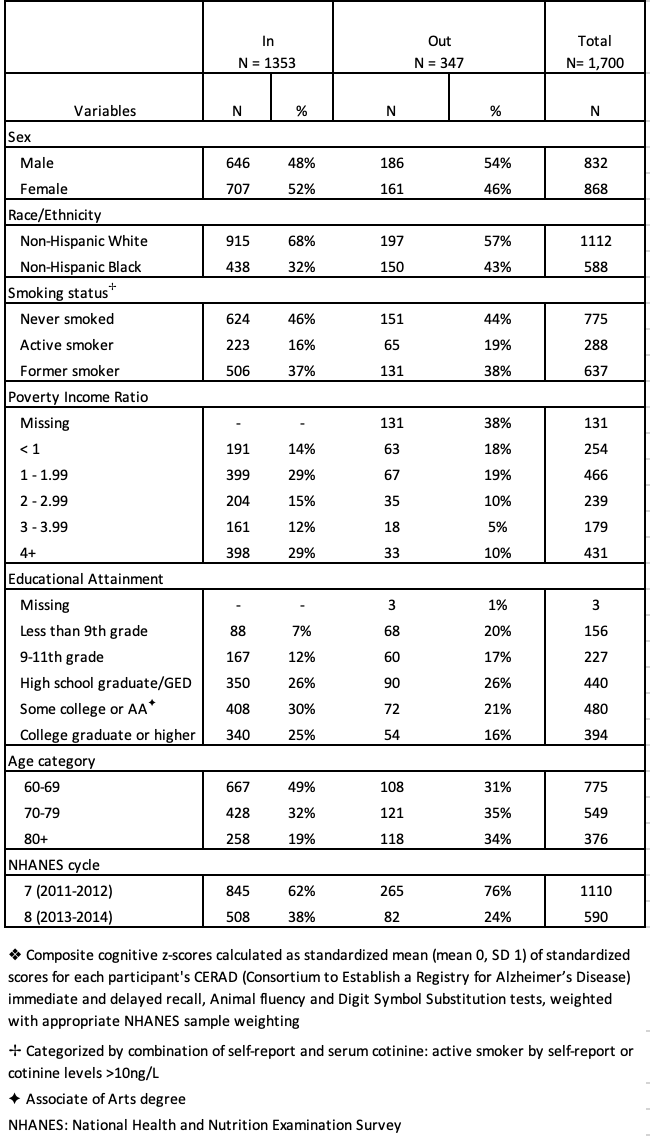

*Web Table 2b.* Covariate missingness for NHANES participants 60+ years and older sub-sampled for blood metals, for composite cognitive z-score analysis (2011-2014, N = 1700).

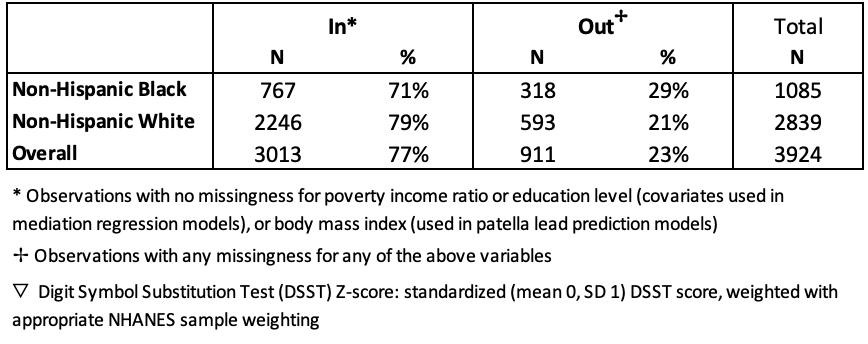

*Web Table 3b*. Covariate missingness for NHANES participants 60+ years and older sub-sampled for blood metals, for Digit Symbol Substitution Test z-score analysis (1999-2002 and 2011-2014, N = 3924)

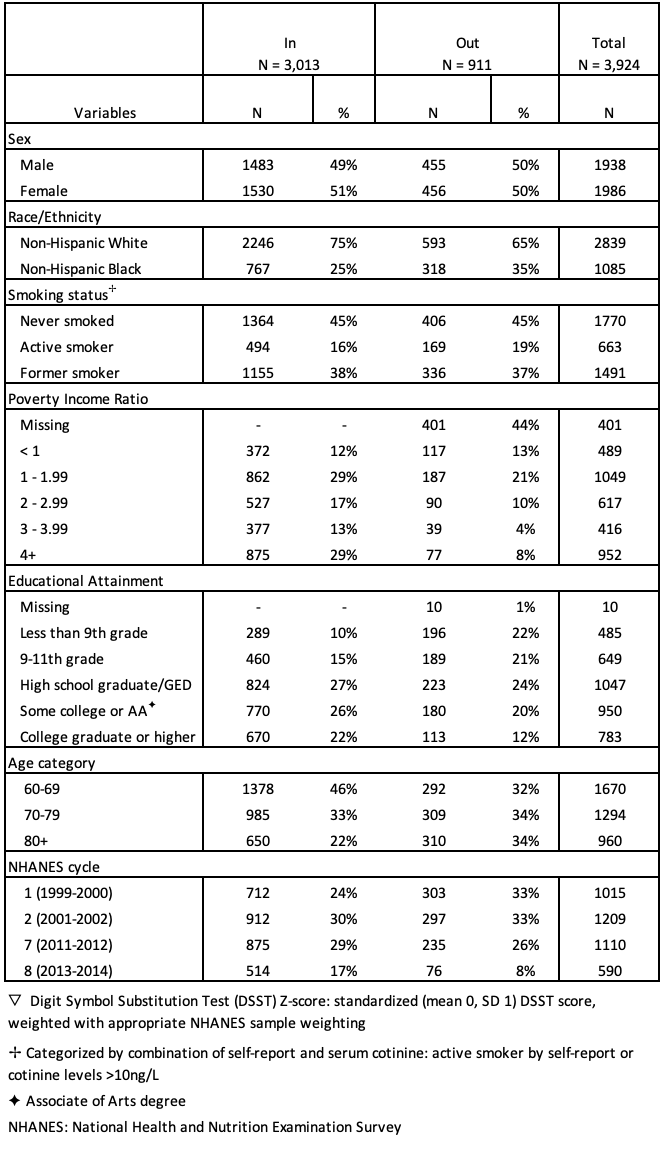

*Web Table 3b.* Covariate missingness for NHANES participants 60+ years and older sub-sampled for blood metals, for Digit Symbol Substitution Test z-score analysis (1999-2002 and 2011-2014, N = 3924)

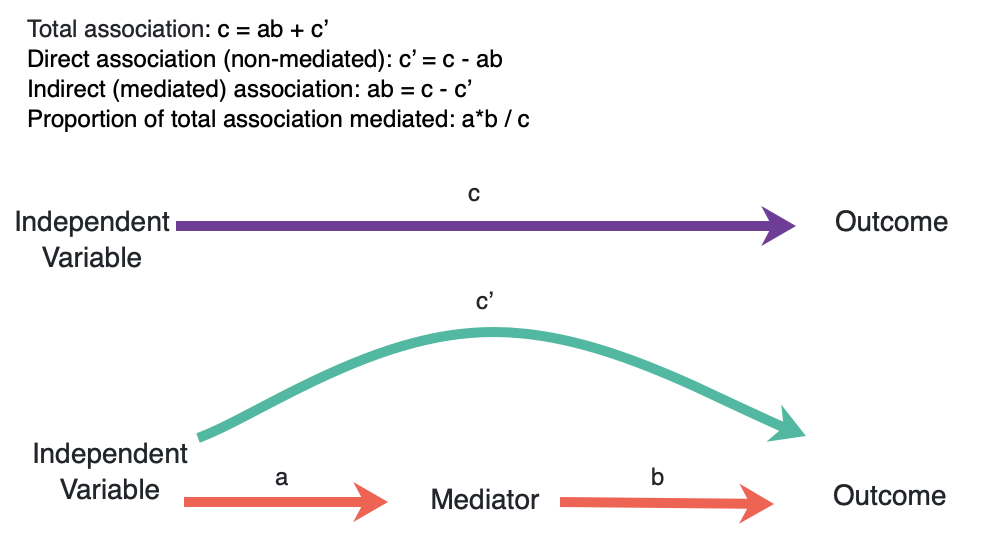

*Web Figure 2*. Baron and Kenny approach for step-by-step estimation of mediated effects

Directed acyclic graph

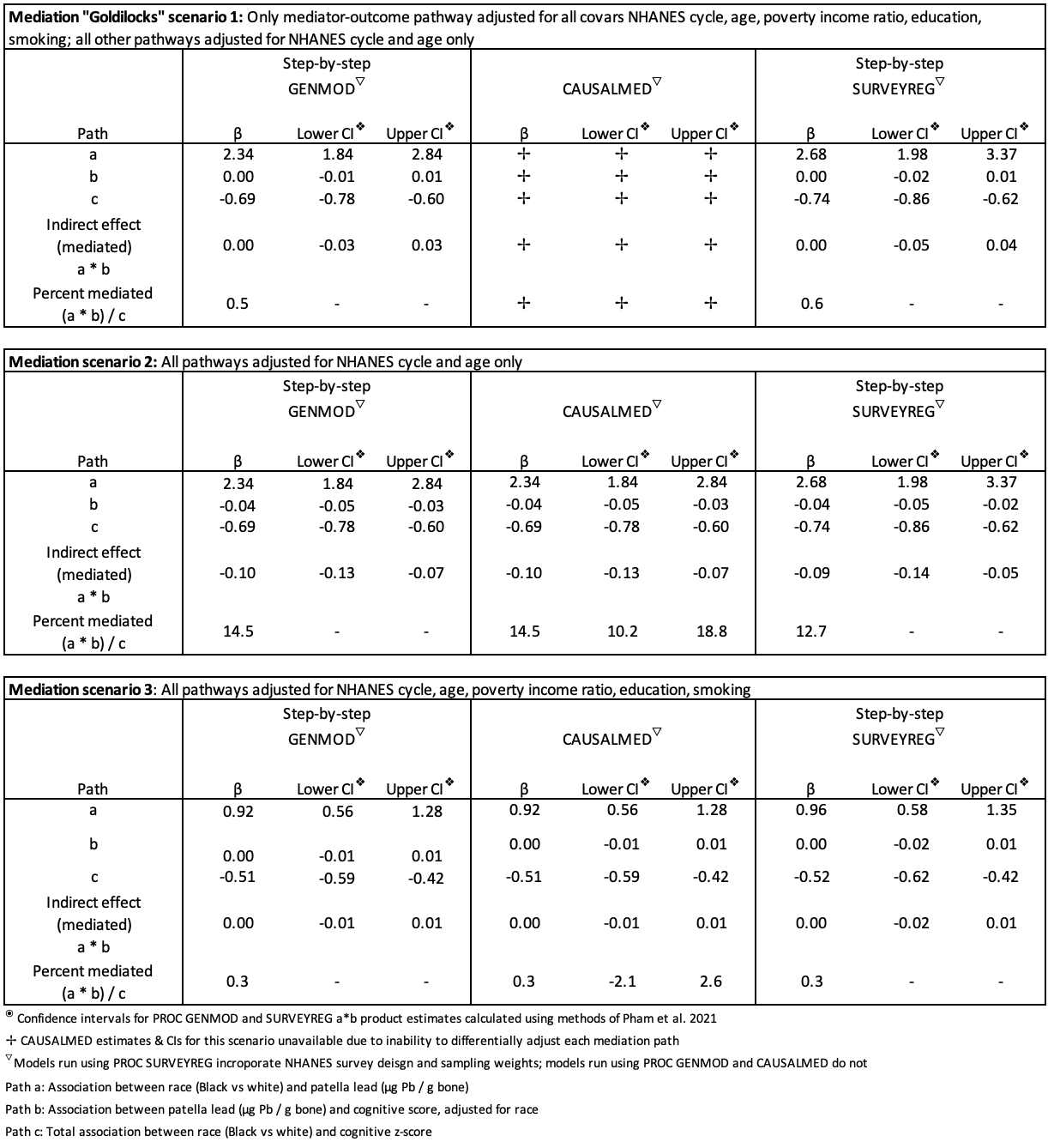

*Web Table 4*. Sensitivity Analyses – mediation results using PROC GENMOD and CAUSALMED for composite cognitive score analyses (NHANES 2011-2014)

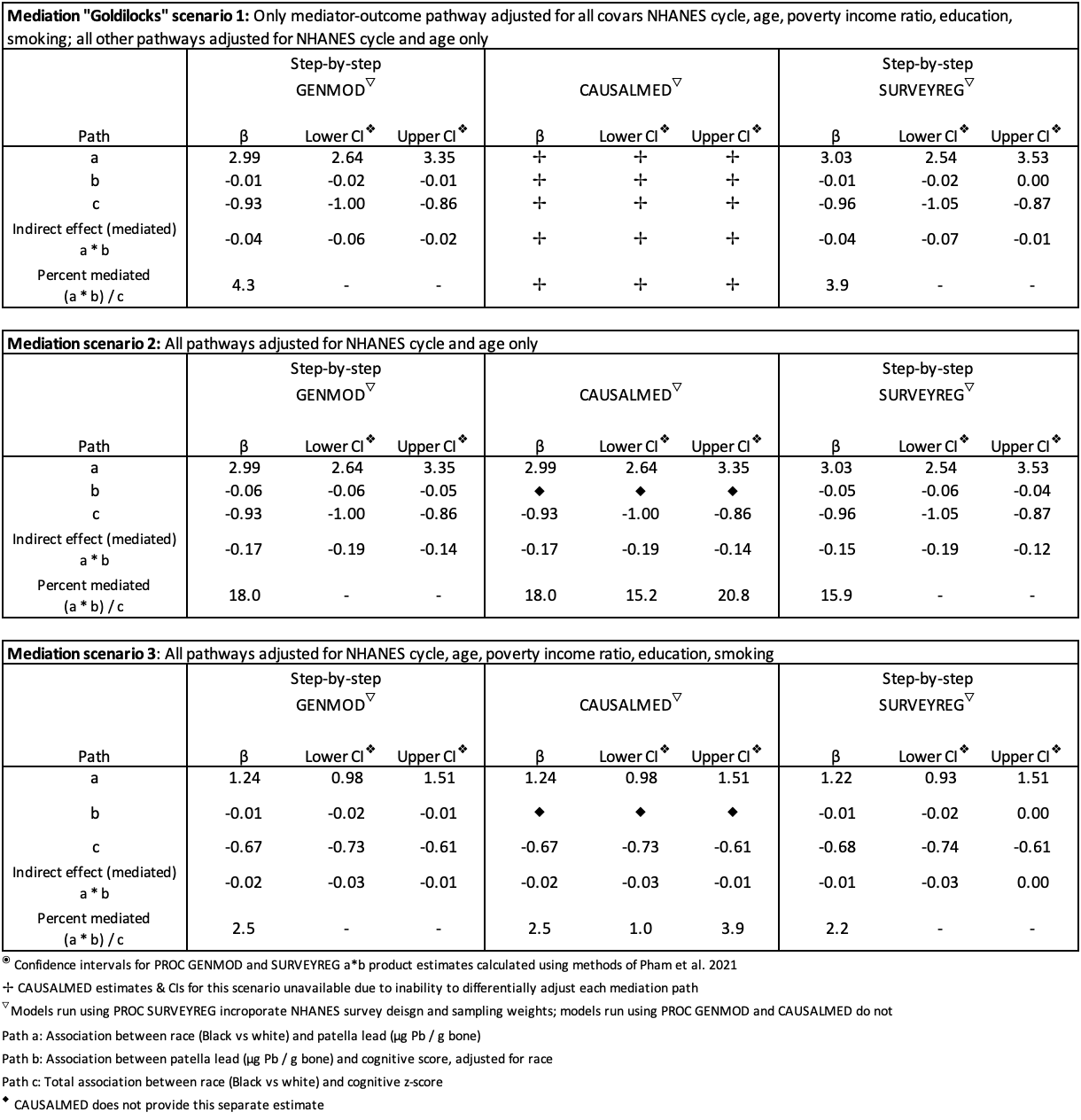

*Web Table 5*. Sensitivity Analyses – mediation results using PROC GENMOD and CAUSALMED for DSST z-score analyses (NHANES 1999-2002 + 2011-2014)
